# Associations of adherence to the World Cancer Research Fund lifestyle recommendations with health-related quality of life and fatigue in Adolescent and Young Adult (AYA) cancer survivors: results from the SURVAYA study

**DOI:** 10.1101/2025.08.22.25334152

**Authors:** Marlou Floor Kenkhuis, Carla Vlooswijk, Silvie H.M. Janssen, Suzanne E. J. Kaal, Jacqueline M. Tromp, Monique E. M. M. Bos, Tom van der Hulle, Roy I. Lalisang, Janine Nuver, Rhodé M. Bijlsma, Winette T.A. van der Graaf, Olga Husson, Laurien M. Buffart

## Abstract

**Purpose:** Adolescents and young adults (AYAs) with cancer face unique long-term social and health challenges that impact their health-related quality of life (HRQoL). This study explores the association between lifestyle behaviors (physical activity, body composition, and nutrition) and HRQoL as well as fatigue in AYA cancer survivors.

**Methods:** The cross-sectional SURVAYA study analyzed data from long-term AYA cancer survivors (5-20 years post diagnosis, aged 18-39 at diagnosis) in The Netherlands. Adherence to the World Cancer Research Fund/American Institute for Cancer Research (WCRF/AICR) lifestyle guidelines was assessed through self-reported questionnaires. HRQoL and fatigue were measured using the EORTC QLQ-C30 questionnaire. Multivariable linear regression was performed to examine associations between lifestyle adherence and HRQoL and fatigue outcomes.

**Results:** The mean adherence to WCRF/AICR recommendations was low to moderate (3.2±1.2, range:0.5-6.0; n=3,668). Adherence rates varied, with 61% of participants limiting the consumption of sugar-sweetened drinks, 28% having a healthy body composition, 25% meeting fruit and vegetable intake recommendations, and 31% limiting alcohol consumption. Better adherence to lifestyle recommendations, particularly in physical activity, healthy body composition, and fruit and vegetable intake, was significantly associated with better physical functioning, global quality of life and less fatigue and pain. Chemotherapy and time since diagnosis were significant moderators of these associations.

**Conclusion:** These findings highlight that better adherence to the WCRF/AICR recommendations is associated with better HRQoL and less fatigue in long-term AYA cancer survivors. Future research should focus on developing targeted interventions to enhance lifestyle adherence and evaluating the long-term effects of these behaviors on survivorship outcomes.

## Introduction

Adolescents and young adults (AYAs) diagnosed with cancer represent a unique but often overlooked patient population in oncology care(1–3). In the Netherlands, AYA cancer patients are defined as patients aged 18 to 39 years at diagnosis, the transition period between centralized pediatric (0-18 years) and non-centralized adult oncology(3). Despite the relatively low incidence compared to older adults, cancer is the second leading cause of disease-related death in AYAs(4). While survival rates for certain cancers in AYAs have improved, late and long-term side effects and elevated risk of developing new malignancies impact health-related quality of life (HRQoL) among AYA cancer survivors(5–7). Fatigue, a well-documented problem during active treatment, often persists for years after therapy(8–10), affecting personal relationships, education, employment, and daily activities(8). Considering their long remaining life span after a cancer diagnosis at a young age, targeting adverse effects of cancer treatment such as fatigue and enhancing HRQoL are imperative.

The 2018 World Cancer Research Fund and the American Institute for Cancer Research (WCRF/AIRC) lifestyle recommendations advocate a healthy lifestyle for cancer survivors of all ages, emphasizing a healthy body composition, regular physical activity (PA), and nutrition including -a diet rich in fruit, vegetables, and dietary fiber, a diet low in red and processed meat, fast food, sugar-sweetened drinks, alcohol- and non-smoking. Adherence to these guidelines is associated with decreased cancer recurrence and improved survival rates among survivors, as well as reduced fatigue and enhanced HRQoL(11–13). However, specific guidelines tailored for AYA cancer survivors remain unavailable.

Numerous studies, including randomized controlled trials, have linked individual lifestyle factors like PA to fatigue and HRQoL in AYA cancer survivors. PA has been shown to have significant beneficial effects on physical fitness and HRQoL(14–16). Similarly, dietary interventions hold promise in improving dietary quality and body composition in AYA cancer survivors, although associations with HRQoL and fatigue remain unclear(17, 18). Recognizing the possible synergistic benefits of the WCRF/AICR lifestyle recommendations, this study examines the association between adherence to the comprehensive set of lifestyle recommendations as a whole, as well as its individual components (body composition, PA and nutrition) and HRQoL and fatigue in a unique population of AYA cancer survivors. These insights aim to inform interventions that enhance long-term well-being in this population.

## Methods

### Design of the study

The study used data from the cross-sectional SURVAYA study, which examined HRQoL and late effects in long-term AYA cancer survivors. It included individuals diagnosed with a primary cancer between ages 18 and 39 from 1999 to 2015, who were 5-20 years post-diagnosis. Participants were selected from the Netherlands Cancer Registry, a population-based registry maintained by the Netherlands Comprehensive Cancer Organisation. Detailed study information is available in Vlooswijk et al.(19). The study adhered to the Declaration of Helsinki, received approval from the Netherlands Cancer Institute Institutional Review Board (IRB-IRBd18122), and was registered in the Clinical trial Registry (NCT05379387). All participants provided written informed consent.

### Data collection

Data collection took place between May 2019 and June 2021 utilizing the Patient Reported Outcomes Following Initial treatment and Long-term Evaluation of Survivorship (PROFILES) system(20), established in 2009. This system, combined with clinical records from the NCR, forms the SURVAYA database. Eligible survivors were invited through their former or current treatment medical practitioners. This communication included comprehensive information about the study and provided AYA cancer survivors with a secure link for accessing log-in instruction, an online informed consent form, and a web-based questionnaire. Additionally, AYA cancer survivors were offered the alternative of receiving a paper version of the questionnaire for completion and return via post. To ensure comprehensive data collection, reminders were dispatched to non-responders within a 2-7-month window. It was explicitly communicated to AYA cancer survivors that their decision to participate or not would not impact their treatment or ongoing care.

### Measures

#### Lifestyle

Adherence to the 2018 WCRF/AICR cancer prevention recommendations was assessed using a standardized scoring system developed by Shams-White et al.(21), excluding alcohol due to inconsistent associations with HRQoL outcomes(22–24). This system served as an indicator of overall lifestyle adherence(25). Six recommendations were operationalized 1) body composition covering BMI and waist circumference, 2) PA, and the nutrition recommendations including 3) fruit and vegetables intake, 4) consumption of ultra-processed foods (in this study defined as cookies, cakes, chips, candy and fast foods), 5) red and processed meat, and 6) sugary drinks. For each recommendation, participants could score 1 (complete adherence), 0.5 (partial adherence) or 0 (non-adherence) points, summed into a total score (range: 0-6), with higher scores indicating better adherence.

PA was assessed using questions adapted from the validated European Prospective Investigation into Cancer (EPIC) Physical Activity Questionnaire(26). Participants reported weekly time (hrs/wk) spent on activities, including walking, cycling, gardening, housekeeping, and sports, for summer and winter separately. To incorporate intensity estimates, metabolic equivalent intensity values (MET) were assigned to each activity based on the compendium of physical activities(27, 28). Moderate-to-vigorous physical activity (MVPA) was defined as activities with MET ≥ 3. The average number of hours per week spent in MVPA was subsequently calculated(28, 29). Given the high adherence rate (98%) to the Dutch PA guideline (2.5 hours of MVPA/wk)(30), participants were categorized into tertiles based on weekly MVPA minutes: the 1^st^ tertile (≤435 min MVPA/wk) representing lower activity levels, and the 3^rd^ tertile (>825 min MVPA/wk) representing higher PA levels.

Body height and weight were self-reported, with BMI classified as underweight (<18.5 kg/m^2^), healthy weight (18.5-24.9 kg/m^2^), overweight (25-29.9 kg/m^2^) and obese (BMI ≥30 kg/m^2^). Waist circumference was self-measured using provided instructions and a measuring tape. Risk cut-offs followed WCRF/AICR, Centers for Disease Control and Prevention(31) and the National Heart, Lung, and Blood Institute guidelines(32): for men <94cm (low risk), between 94 and 102cm (increased risk) and ≥102cm (substantially increased risk); for women, <80cm (low risk), between 80 and 88 cm (increased risk) and ≥88 cm (substantially increased risk). Body composition score was based on both the BMI categorization and waist circumference categorization, where BMI and waist circumference each receive up to half a point(21).

Nutrition was evaluated using a 10-item self-administered questionnaire covering food groups such as ‘Vegetables’, ‘Fruit’, ‘Cookies, cakes, chips and candy’, ‘Red meat’, ‘Processed meat’, ‘Sweetened drinks’, and ‘Fast foods’. Participants reported consumption frequency on an eight-point scale (from ‘never’ to ‘more than seven times a week’). For ‘Vegetables’, ‘Fruit’ and ‘Sweetened drinks’, respondents also specified the number of portions consumed daily (from ‘1 to 7 portions per day’ or ‘not applicable’. A portion size was defined as a spoon of vegetables (50g), a piece of fruit (125g), or a glass of sweetened drinks (250ml). The daily consumption of these items was calculated by converting portions to grams and multiplying by the frequency of consumption for each food item. Smoking and alcohol use were assessed through self-developed questions, with alcohol intake recorded as weekly glasses consumed.

#### Patient-reported outcomes

Patient-reported outcomes were measured through the 30-item European Organization for the Research and Treatment of Cancer Quality of Life Questionnaire-Core 30 (EORTC QLQ-C30), which includes five functioning scales (physical, role, cognitive, emotional and social functioning), three symptom scales (fatigue, pain, and nausea and vomiting), a global health/QoL scale and a number of single items for specific cancer-/treatment-related symptoms. Scores range from 0 to 100, with higher scores on the global QoL and functioning scales reflecting better HRQoL or functioning, whereas higher symptom scales indicate more symptoms (i.e. worse fatigue). This study focused on the functioning scales, symptom scales and the summary score of the EORTC QLQ-C30, excluding single-item measures and the nausea symptoms scale, as the patients were post-treatment.

#### Co-variables

Socio-demographic information including sex and age, along with clinical characteristics such as tumor type, cancer stage, primary treatment and date of diagnosis were extracted from the Netherlands Cancer Registry. Tumor types were categorized according to the third International Classification of Diseases for Oncology (ICDO-3)(33). Cancer stage was determined based on the Tumor-Node-Metastasis (TNM) classification system or Ann Arbor Code for Hodgkin lymphoma and Non-Hodgkin lymphoma cases(34). Educational level was self-reported and dichotomized into lower (secondary school or less) and higher (college or university).

#### Statistical analyses

Descriptive statistics (means and standard deviations or frequencies and percentages) were calculated to describe main sample characteristics. To assess the association between the WCRF adherence score (continuous), and the individual 2018 WCRF/AICR recommendations with HRQoL and fatigue, we utilized confounder-adjusted linear regression models. Each individual recommendation was evaluated based on the score of the WCRF/AICR recommendation. HRQoL and fatigue were considered as outcome variables. A set of predefined confounders was included in the analyses: age (years), sex, number of co-morbidities (0,1, ≥2), current smoking (yes, no), educational level (lower, higher), tumor stages (1/2 and 3/4), chemotherapy (yes, no), radiotherapy (yes, no), second tumor (yes, no) and time since diagnosis (years). Results were reported as regression coefficients and 95% confidence intervals (CIs). Potential effect modification (i.e., moderators) of sex, education, comorbidities, chemotherapy and time since diagnoses was explored by including interaction terms in linear regression models. Given the exploratory nature of these analyses, stratified analyses per moderator subgroups were conducted when the interaction term had a p-value less than 0.10. Statistical analyses were conducted using IBM SPSS Statistics (version 29) with statistical significance set at P<0.05 (two-tailed).

## Results

### General characteristics of the study population

A total of 11,340 AYA cancer survivors were invited to participate in the study, of whom 4,010 (35%) completed the questionnaire. Participants with missing data on the WCRF/AICR adherence score (n=342) were excluded, leaving 3,668 AYA cancer survivors for the analysis. The demographic and lifestyle characteristics of these AYA cancer survivors are summarized in Table 1. AYA cancer survivors were mostly women (60.7%), with a mean age of 44.5 ± 7.5 years at the time of completing the questionnaire. On average, participants had been diagnosed 12.4 ± 4.5 years prior to filling out the questionnaire.

**Table 1.**
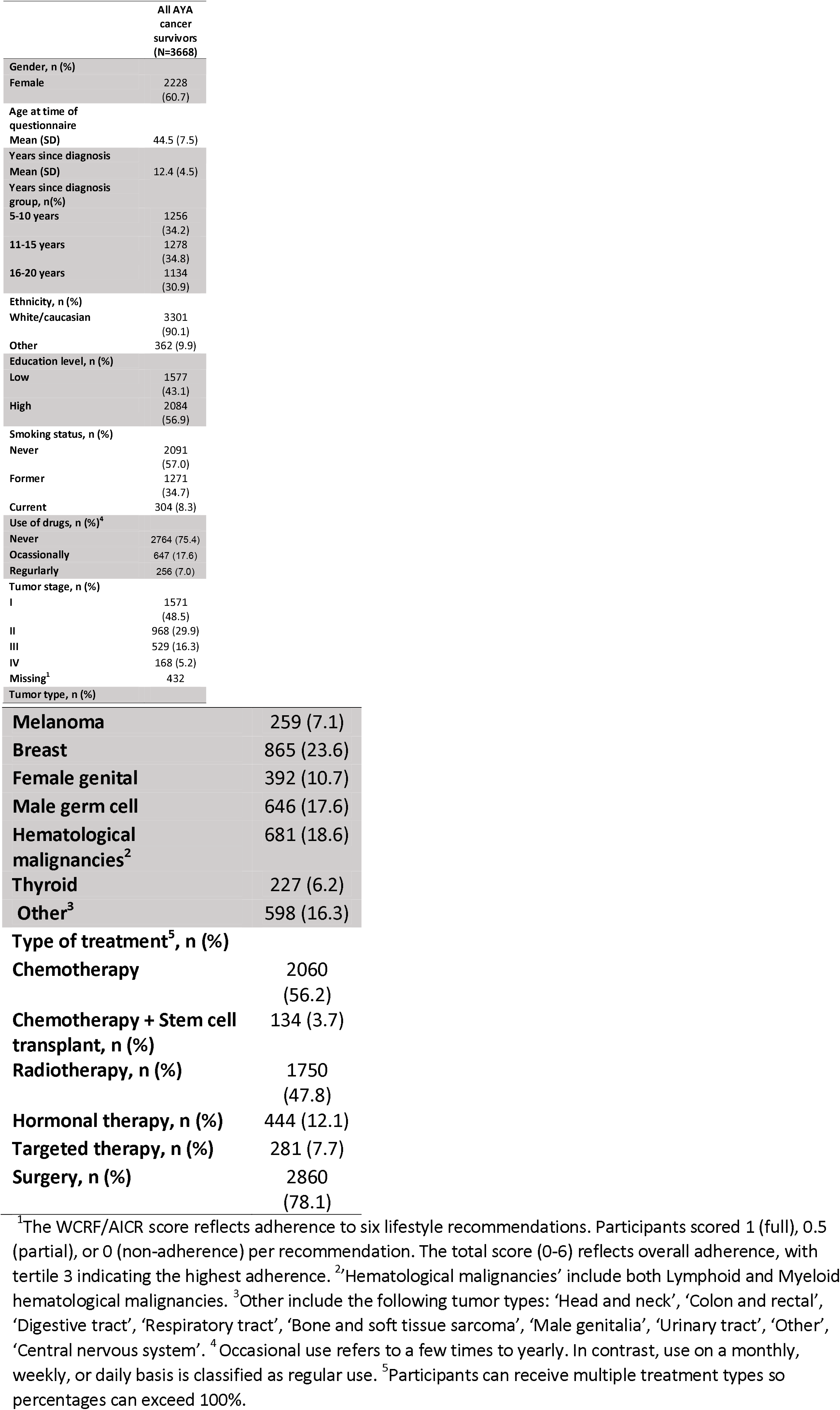
Sociodemographic, and clinical characteristics of AYA cancer survivors.

|  | **All AYA**<br>**cancer**<br>**survivors**<br>**(N=3668)** |
| --- | --- |
| **Gender, n (%)** |  |
| Female | 2228<br>(60.7) |
| **Age at time of**<br>**questionnaire** |  |
| Mean (SD) | 44.5 (7.5) |
| **Years since diagnosis** |  |
| Mean (SD) | 12.4 (4.5) |
| **Years since diagnosis**<br>**group, n(%)** |  |
| 5-10 years | 1256<br>(34.2) |
| 11-15 years | 1278<br>(34.8) |
| 16-20 years | 1134<br>(30.9) |
| **Ethnicity, n (%)** |  |
| White/caucasian | 3301<br>(90.1) |
| Other | 362 (9.9) |
| **Education level, n (%)** |  |
| Low | 1577<br>(43.1) |
| High | 2084<br>(56.9) |
| **Smoking status, n (%)** |  |
| Never | 2091<br>(57.0) |
| Former | 1271<br>(34.7) |
| Current | 304 (8.3) |
| **Use of drugs, n (%)<sup>4</sup>** |  |
| Never | 2764 (75.4) |
| Ocassionally | 647 (17.6) |
| Regurlarly | 256 (7.0) |
| **Tumor stage, n (%)** |  |
| I | 1571<br>(48.5) |
| II | 968 (29.9) |
| III | 529 (16.3) |
| IV | 168 (5.2) |
| Missing<sup>1</sup> | 432 |
| **Tumor type, n (%)** |  |
| **Melanoma** | 259 (7.1) |
| **Breast** | 865 (23.6) |
| **Female genital** | 392 (10.7) |
| **Male germ cell** | 646 (17.6) |
| **Hematological malignancies<sup>2</sup>** | 681 (18.6) |
| **Thyroid** | 227 (6.2) |
| **Other<sup>3</sup>** | 598 (16.3) |
| **Type of treatment<sup>5</sup>, n (%)** |  |
| **Chemotherapy** | 2060<br>(56.2) |
| **Chemotherapy + Stem cell transplant, n (%)** | 134 (3.7) |
| **Radiotherapy, n (%)** | 1750<br>(47.8) |
| **Hormonal therapy, n (%)** | 444 (12.1) |
| **Targeted therapy, n (%)** | 281 (7.7) |
| **Surgery, n (%)** | 2860<br>(78.1) |

<sup>1</sup>The WCRF/AICR score reflects adherence to six lifestyle recommendations. Participants scored 1 (full), 0.5 (partial), or 0 (non-adherence) per recommendation. The total score (0-6) reflects overall adherence, with tertile 3 indicating the highest adherence. <sup>2</sup>'Hematological malignancies' include both Lymphoid and Myeloid hematological malignancies. <sup>3</sup>Other include the following tumor types: 'Head and neck', 'Colon and rectal', 'Digestive tract', 'Respiratory tract', 'Bone and soft tissue sarcoma', 'Male genitalia', 'Urinary tract', 'Other', 'Central nervous system'. <sup>4</sup>Occasional use refers to a few times to yearly. In contrast, use on a monthly, weekly, or daily basis is classified as regular use. <sup>5</sup>Participants can receive multiple treatment types so percentages can exceed 100%.

**Table 1.** Cross-sectional associations (regression coefficients and 95% confidence intervals) between the WCRF/AICR lifestyle recommendations score and.

|  |  | Physical<br>Functioning | Role<br>Functioning | Emotional<br>Functioning | Cognitive<br>Functioning | Social<br>Functioning | Global<br>Quality of<br>Life | Fatigue | Pain | EORTC<br>Summary<br>Score |
| --- | --- | --- | --- | --- | --- | --- | --- | --- | --- | --- |
| WCRF Score<br>(continuous) |  | 1.0*<br>(0.6; 1.4) | 0.8*<br>(0.1; 1.6) | 0.8*<br>(0.2; 1.4) | 0.4<br>(-0.3; 1.1) | 0.6<br>(-0.1; 1.2) | 1.6*<br>(1.1; 2.1) | -1.1*<br>(-1.8; -0.4) | -1.1*<br>(-1.7; -0.4) | 0.5*<br>(0.1; 0.8) |
| Physical activity<br>Score | 0 | REF | REF | REF | REF | REF | REF | REF | REF | REF |
|  | 0.5 | 2.8*<br>(1.7; 3.9) | 2.0<br>(-0.0; 4.0) | 1.5<br>(-0.2; 3.2) | 0.6<br>(-1.4; 2.6) | 0.5<br>(-1.2; 2.3) | 2.8*<br>(1.4; 4.3) | -3.0*<br>(-5.1; -0.9) | -2.3*<br>(-4.1; -0.4) | 1.5*<br>(0.5; 2.5) |
|  | 1<sup>1</sup> | 4.0*<br>(2.9; 5.0) | 3.2*<br>(1.2; 5.2) | 2.0*<br>(0.3; 3.7) | 1.5<br>(-0.5; 3.5) | 1.6<br>(-0.1; 3.4) | 4.4*<br>(2.9; 5.8) | -4.3*<br>(-6.4; -2.2) | -2.4*<br>(-4.2; -0.5) | 1.5*<br>(0.5; 2.5) |
| Body composition<br>Score | 0 | REF | REF | REF | REF | REF | REF | REF | REF | REF |
|  | 0.5 | 2.5*<br>(1.4; 3.5) | 2.8*<br>(0.9; 4.8) | 1.3*<br>(-0.4; 2.9) | 0.8<br>(-1.1; 2.7) | 1.6<br>(-0.1; 3.3) | 2.7*<br>(1.3; 4.1) | -3.5*<br>(-5.5; -1.4) | -3.9*<br>(-5.7; -2.1) | 1.8*<br>(0.8; 2.7) |
|  | 1 | 2.9*<br>(1.7; 4.1) | 3.1*<br>(0.9; 5.3) | 1.2*<br>(-0.6; 3.1) | 1.3<br>(-0.8; 3.4) | 2.1<br>(0.2; 4.0) | 2.8*<br>(1.2; 4.4) | -2.1<br>(-4.3; 0.2) | -4.4*<br>(-6.5; -2.4) | 1.6*<br>(0.5; 2.7) |
| Fruit and vegetable<br>Score | 0 | REF | REF | REF | REF | REF | REF | REF | REF | REF |
|  | 0.5 | 1.4*<br>(0.4; 2.5) | 1.7<br>(-0.2; 3.7) | 0.7<br>(-1.0; 2.4) | 0.7<br>(-1.2; 2.6) | 0.8<br>(-0.9; 2.5) | 1.6*<br>(0.2; 3.0) | -1.8<br>(-3.9; 0.2) | -1.4<br>(-3.3; 0.4) | 1.3*<br>(0.3; 2.2) |
|  | 1 | 1.9*<br>(0.6; 3.1) | 0.8<br>(-1.5; 3.1) | 1.8<br>(-0.2; 3.7) | 2.5*<br>(0.3; 4.8) | 1.5<br>(-0.5; 3.5) | 4.1*<br>(2.5; 5.8) | -2.8*<br>(-5.2; -0.4) | -1.6<br>(-3.7; 0.6) | 1.9*<br>(0.8; 3.1) |
| Sugar-sweetened<br>drinks Score | 0 | REF | REF | REF | REF | REF | REF | REF | REF | REF |
|  | 0.5 | 0.2<br>(-1.4; 1.8) | 2.9<br>(-0.1; 5.9) | 2.4<br>(-0.1; 4.9) | 0.4<br>(-2.5; 3.3) | 1.5<br>(-1.1; 4.1) | 2.1<br>(-0.1; 4.2) | -4.6*<br>(-7.6; -1.5) | -3.3*<br>(-6.0; -0.6) | 1.5*<br>(0.0; 3.0) |
|  | 1 | 1.5<br>(-0.0; 3.0) | 4.0*<br>(1.2; 6.8) | 2.7*<br>(0.4; 5.1) | 1.6<br>(-1.1; 4.4) | 2.6*<br>(0.1; 5.0) | 3.1*<br>(1.1; 5.1) | -4.7*<br>(-7.6; -1.8) | -4.1*<br>(-6.7; -1.5) | 2.2*<br>(0.8; 3.6) |
| Ultra processed foods<br>Score | 0 | REF | REF | REF | REF | REF | REF | REF | REF | REF |
|  | 0.5 | 0.5<br>(-0.6; 1.6) | 0.9<br>(-1.1; 2.9) | 1.8*<br>(0.1; 3.5) | 0.7<br>(-1.2; 2.7) | 0.6<br>(-1.1; 2.3) | 1.4<br>(-0.0; 2.9) | -0.8<br>(-2.8; 1.3) | -0.8<br>(-2.6; 1.1) | 0.6<br>(-0.4; 1.6) |
|  | 1 | -0.9 | -1.0 | 2.0* | -0.8 | -0.5 | 0.7 | -0.2 | 0.5 | -0.4 |
|  |  | (-1.9; 0.2) | (-3.0; 0.9) | (0.4; 3.7) | (-2.7; 1.1) | (-2.2; 1.2) | (-0.7; 2.1) | (-2.2; 1.9) | (-1.3; 2.3) | (-1.4; 0.6) |
| **Red and processed meat Score** | 0 | REF | REF | REF | REF | REF | REF | REF | REF | REF |
|  | 0.5 | -1.1* | -0.8 | -1.6 | -1.0 | -0.4 | -0.9 | 3.0* | 1.4 | -1.3* |
|  |  | (-2.1; -0.0) | (-2.7; 1.1) | (-3.2; 0.0) | (-2.8; 0.9) | (-2.0; 1.3) | (-2.3; 0.5) | (1.0; 5.0) | (-0.4; 3.2) | (-2.2; -0.3) |
|  | 1 | -0.5 | -2.1 | -2.4 | -2.0 | -1.9* | -0.5 | 2.9* | 0.8 | -1.9* |
|  |  | (-1.6; 0.7) | (-4.2; 0.0) | (-4.1; -0.6) | (-4.1; 0.0) | (-3.8; -0.1) | (-2.0; 1.0) | (0.7; 5.1) | (-1.1; 2.8) | (-2.9; -0.8) |
| **Alcohol Score** | 0 | REF | REF | REF | REF | REF | REF | REF | REF | REF |
|  | 0.5 | -0.7 | -0.5 | 1.7 | 0.8 | 0.0 | 2.3 | -1.5 | -1.8 | 0.5 |
|  |  | (-2.4; 1.1) | (-3.7; 2.7) | (-1.1; 4.4) | (-2.4; 4.0) | (-2.9; 2.8) | (-0.1; 4.6) | (-4.9; 1.9) | (-4.9; 1.2) | (-1.2; 2.1) |
|  | 1 | -6.3* | -10.3* | -2.9* | -3.8* | -7.8* | -3.5* | 5.7* | 4.8* | -3.8* |
|  |  | (-9.8; -4.5) | (-13.7; -6.9) | (-5.8; -0.0) | (-7.2; -0.5) | (-10.8; -4.8) | (-5.9; -1.0) | (2.1; 9.2) | (1.6; 7.9) | (-5.5; -2.1) |

<sup>1</sup>Scores represent level of adherence: 0 (non-adherence), 0.5 (partial adherence), 1 (full adherence). \* Statistically significant
*Note:* All analyses were adjusted for age (years), sex, number of co-morbidities (0,1, ≥2), current smoking (yes, no), educational level (lower, higher), tumor stages (1/2 and 3/4), chemotherapy (yes, no), radiotherapy (yes, no), second tumor (yes, no) and time since diagnosis (years).

**Table 1.** Moderator effects and stratified analysis of the association between the WCRF/AICR lifestyle recommendations score with HRQoL outcomes and gue in AYA cancer survivors (n=3668).

| Moderators | Physical Functioning | Role Functioning | Emotional Functioning | Cognitive Functioning | Social Functioning | Global Quality of Life | Fatigue | Pain | EORTC Summary Score |
| --- | --- | --- | --- | --- | --- | --- | --- | --- | --- |
| **Sex<sup>a</sup>** | 0.06* | 0.83 | 0.90 | 0.65 | 0.67 | 0.07* | 0.78 | 0.96 | 0.61 |
| **Male<sup>b</sup>** | 0.5 (-0.0; 1.1) | Na | Na | Na | Na | 2.0 (1.2; 2.8)* | Na | Na | Na |
| **Female<sup>b</sup>** | 1.3 (0.8; 1.8)* | Na | Na | Na | Na | 1.3 (0.6;1.9)* | Na | Na | Na |
| **Education<sup>a</sup>** | 0.76 | 0.81 | 0.36 | 0.48 | 0.90 | 0.31 | 0.24 | 0.34 | 0.17 |
| **Comorbidities<sup>a</sup>** | 0.02* | 0.48 | 0.24 | 0.44 | 0.85 | 0.12 | 0.94 | 0.71 | 0.20 |
| **0<sup>b</sup>** | 0.8 (0.4; 1.2)* | Na | Na | Na | Na | Na | Na | Na | Na |
| **≥1<sup>b</sup>** | 1.5 (0.6; 2.5)* | Na | Na | Na | Na | Na | Na | Na | Na |
| **Chemotherapy<sup>a</sup>** | 0.06* | 0.03* | 0.03* | 0.64 | 0.01* | 0.02* | 0.06* | <0.00* | 0.02* |
| **Yes<sup>b</sup>** | 0.7 (0.2; 1.2)* | 0.2 (-0.7; 1.2) | 0.3 (-0.5; 1.2) | Na | -0.1 (-1.0; 0.8) | 1.1 (0.4; 1.8) | -0.7 (-1.7; 0.3) | -0.2 (-1.1; 0.7) | 0.2 (-0.3; 0.7) |
| **No<sup>b</sup>** | 1.4 (0.8; 2.0)* | 1.6 (0.5; 2.7)* | 1.4 (0.5; 2.3)* | Na | 1.4 (0.6; 2.3)* | 2.2 (1.4; 2.9)* | -1.6 (-2.7; -0.5)* | -2.2 (-3.2; -1.3)* | 0.9 (0.4; 1.4)* |
| **Tumor stage<sup>a</sup>** | 0.87 | 0.49 | 0.02 | 0.02 | 0.02 | 0.17 | 0.26 | 0.81 | 0.32 |
| **Low (I/II)<sup>b</sup>** | Na | Na | 1.2 (0.5; 1.8)* | 0.8 (0.0; 1.5)* | 0.9 (0.2; 1.6)* | Na | Na | Na | Na |
| **High (III/IV)<sup>b</sup>** | Na | Na | -0.7 (-2.1; 0.6) | -1.3 (-3.0; 0.3) | -0.8 (-2.3; 0.7) | Na | Na | Na | Na |
| **Time since diagnoses<sup>a</sup>** | 0.09* | 0.23 | 0.10 | 0.19 | 0.40 | 0.57 | 0.05 | <0.001* | 0.02* |
| **5-10 years<sup>b</sup>** | 0.5 (-0.2; 1.2) | Na | Na | Na | Na | Na | -0.03 (-1.4; 1.3) | 0.4 (-0.8; 1.5) | -0.1 (-0.8; 0.6) |
| **11-15 years<sup>b</sup>** | 1.0 (0.4; 1.7)* | Na | Na | Na | Na | Na | -1.6 (-2.8; -0.4)* | -1.4 (-2.5; -0.3)* | 0.5 (-0.0; 1.1) |
| **16-20 years<sup>b</sup>** | 1.6 (0.9; 2.3)* | Na | Na | Na | Na | Na | -1.5 (-2.8; -0.2)* | -2.1 (-2.8; -0.2)* | 1.0 (0.3; 1.6)* |

\*Statistically significant p<0.05
<sup>a</sup>P-value for interaction
<sup>b</sup>Beta and 95% confidence interval

### WCRF/AICR adherence score

The mean total WCRF/AICR adherence score was 3.2 ± 1.2, with scores ranging from 0.0 to 6.0. Adherence to the individual WCRF/AICR recommendations varied among participants. Sixty-one percent fully adhered to the recommendation to limit the consumption of sugar-sweetened drinks, 28% met the recommendation for a healthy body composition, 25% met the recommendations for fruit and vegetable consumption and 31% adhered to the recommendation to limit alcohol consumption (Figure 1).

**Figure 1.**
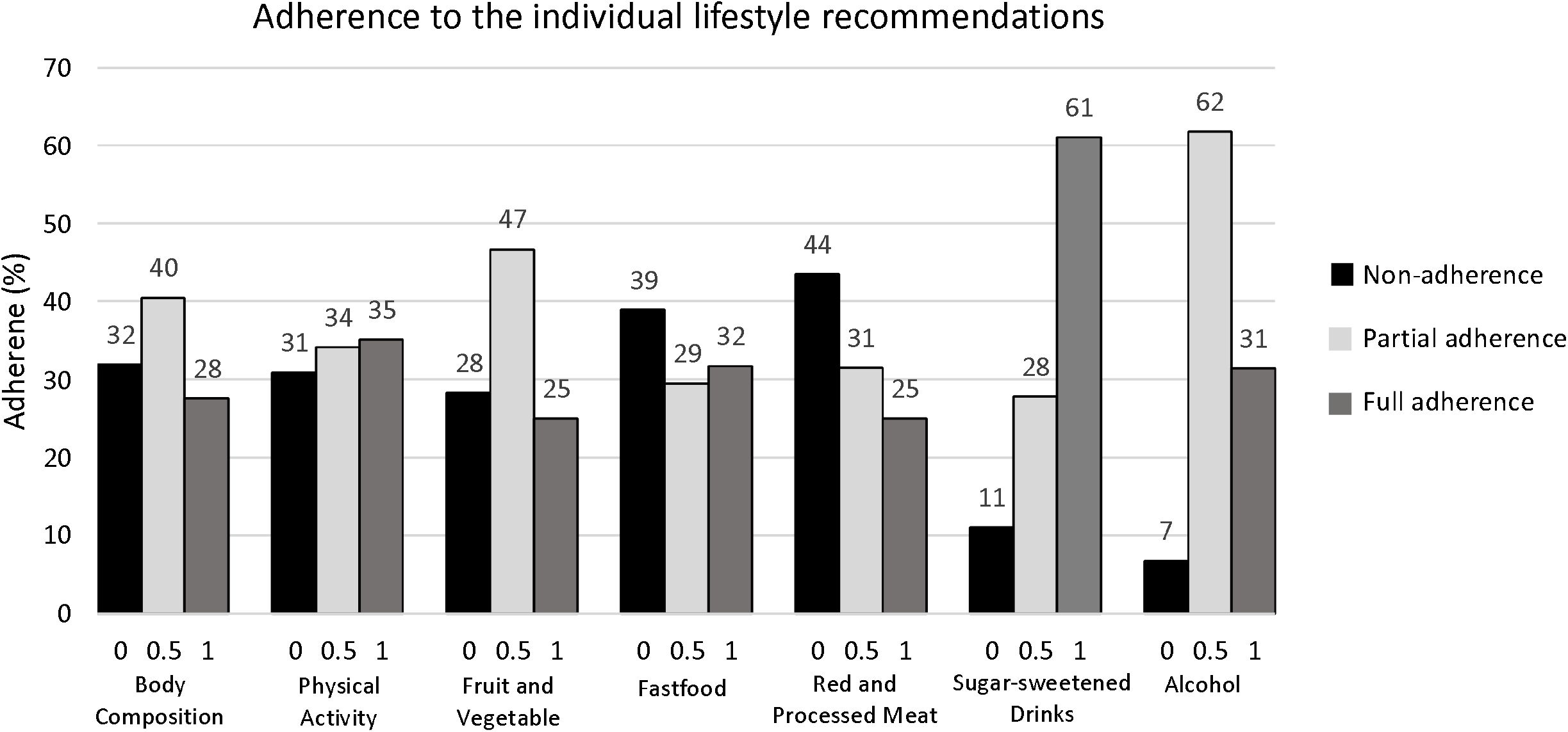
Adherence to the individual World Cancer Research Fund and American Institute for Cancer Research lifestyle recommendations. Scores (x-as) represent level of adherence: 0 (non-adherence), 0.5 (partial adherence), 1 (full adherence) and were based on cut-off values reported in the standardized scoring system developed by Shams-White et al. (28)

### Lifestyle associations with HRQoL and fatigue

In confounder-adjusted multivariable linear regression models, better adherence to the WCRF/AICR guidelines was significantly associated with better physical functioning (β:1.0; 95%CI: 0.6; 1.4), role functioning (β:0.8; 95%CI: 0.1; 1.6), emotional functioning (β:0.8; 95%CI: 0.2; 1.4), global quality of life (β:1.6; 95%CI: 1.1; 2.1), and EORTC summary score (β:0.5; 95%CI: 0.1; 0.8), but also with less fatigue (β:−1.1; 95%CI: −1.8; −0.4) and pain symptoms (β:−1.1; 95%CI: −1.7; 0.4) (Table 2).

Chemotherapy moderated the associations between adherence to the WCRF guidelines for all outcomes except for cognitive functioning. Stratified analyses revealed that these associations were statistically significant in participants who did not receive chemotherapy but not in) those who did receive chemotherapy (Table 3). In addition, tumor stage moderated the association between adherence to the WCRF guidelines for emotional functioning (p=0.02), cognitive functioning (p=0.02), and social functioning (p=0.02), with statistically significant associations among participants who had a low stage (stage I/II) but not in those with a high stage (III/IV). Lastly, time since diagnosis moderated the associations for physical functioning (p=0.09), fatigue (p=0.05), pain (p<0.01) and the EORTC summary score (p=0.02), with stronger associations in participants who were 16-20 years post-diagnosis compared to those 11-15 years or those 5-10 years post-diagnosis.

Results for the individual recommendations showed that better adherence to the recommendations of PA, body composition, fruit and vegetable consumption and reduced intake of sugar sweetened drink were significantly associated with better physical functioning, global quality of life, and the EORTC summary score, and with less fatigue. Conversely, a negative association was found for use of alcohol, where participants adhering to this recommendation (e.g. limiting alcohol consumption) experienced lower physical functioning, global quality of life, and more fatigue (Table 2).

## Discussion

In this population-based, cross-sectional study among AYA cancer survivors, we found low-moderate overall adherence to the 2018 WCRF/AICR recommendations, particularly for having a healthy body composition and the consumption of fruit and vegetables. Overall healthy lifestyle was associated with better functioning on all HRQoL domains, except for cognitive functioning, and less fatigue and pain. This association was especially evident for the individual lifestyle components of PA, body composition and fruit and vegetable intake and not for sugar-sweetened drinks, ultra-processed foods and red and processed meat.

### Adherence to WCRF/AICR recommendations

The low to moderate adherence among AYA cancer survivors, especially regarding a healthy body composition and consumption of fruits, vegetables and alcohol, mirrors findings seen in other adult populations of cancer survivors, such as those with colorectal or bladder cancer(35–37), as well as the general population(38). This underscores the broader challenges of adopting and maintaining a healthy lifestyle, both in general and after cancer treatment.

### Associations between lifestyle and HRQoL and fatigue

The global quality of life of AYA cancer survivors appears comparable to normative AYA populations(39), nevertheless our findings indicate a positive association between better lifestyle adherence and better HRQoL and less fatigue and pain. This underscores the potential benefits of promoting health behaviors in this group, particularly in areas of low adherence such as body composition and fruit and vegetable intake, as well as PA, which shows the strongest associations. Although the overall association between adherence to the WCRF guidelines and cognitive functioning was limited, our findings suggest that tumor stage moderated this association. This may be attributed to the increased risk of cognitive impairment linked to various factors such as treatment type, intensity, and duration(40), which tend to be more pronounced in higher tumor stages. These factors may attenuate or diminish the potential benefits of lifestyle on cognitive functioning. Consequently, better lifestyle adherence may positively influence cognitive functioning in patients diagnosed at earlier stages of cancer. In contrast, addressing cognitive functioning in survivors with more advanced disease may require alternative or more targeted approaches beyond lifestyle modification, including multimodal or specialized cognitive rehabilitation strategies (41). The distinction between the different HRQoL domains, such as physical and cognitive functioning, underscores the need for differentiated intervention strategies. Tailoring support to the specific challenges faced by AYA cancer survivors is essential for addressing their diverse needs (42). Lastly, the alcohol score in this paper showed similar results to what has been previously described showing lower HRQoL and more fatigue and pain in participants who adhere to the alcohol recommendation and do not drink. This may reflect that participants who feel better and less fatigued in the post-treatment period may be more likely to consume alcohol, as feeling better may lead to more socializing and participation in activities commonly associated with alcohol.

Additionally, chemotherapy emerged as a moderating factor, with stronger associations observed among AYA cancer survivors who did not receive chemotherapy compared to those who did as supported by a study in breast cancer survivors(43). The lasting side effects of chemotherapy may reduce the impact of health behaviors on HRQoL, indicating that survivors who received chemotherapy may need more targeted or intensive support to address treatment-specific challenges that lifestyle changes alone cannot fully address. Furthermore, the perception of treatment-related side effects may also influence HRQoL(44). Survivors who believe chemotherapy will lead to long-term physical limitations may perceive their HRQoL as lower. Several studies have indicated that treatment perceptions can affect HRQoL(44–46).

Moreover, the association between lifestyle adherence and HRQoL was more pronounced in survivors 15-20 years post-diagnosis compared to those 5-10 or 10-15 years post-diagnosis. This may suggest that the benefits of lifestyle adherence may accumulate over time or become more apparent as survivors age. These findings emphasize the importance of sustained health behavior promotion and follow-up, particularly as survivors move further from their treatment. Early-stage survivors might benefit more from interventions aimed at addressing immediate post-treatment challenges, while later-stage survivors may respond to strategies emphasizing long-term health promotion.

### Strengths and limitations

Strengths of this study include its large sample size, population-based approach, and extended post-diagnosis timeline. However, the cross-sectional design limits our ability to establish causal relationships and to determine whether adherence to recommendations reflects long-term behavioral patterns. Additionally, the use of self-reported data introduces potential biases. Self-reported measures of body weight may have led to an underestimation of BMI, especially in overweight and obese individuals, thereby potentially even lowering the prevalence within the healthy BMI category(47). Additionally, while 98% of participants appeared to adhere to the PA guidelines, this proportion is likely overestimated due to the use of self-reports to assess PA. The use of a shortened version of the EPIC PAQ may have contributed to inaccuracies by limiting the ability to assess the intensity of PA. Participants may have overestimated leisure activities and the intensity of sports activities. Cultural factors, such as high levels of cycling for daily purposes instead of sports among Dutch participants, may have also inflated adherence estimates, both in terms of duration and intensity(48). Therefore, we decided to categorize our MVPA data in tertiles to distinguish between more and less active AYA cancer survivors and to limit potential bias in the associations. These examples underscore the challenges of interpreting self-reported lifestyle behaviors and these biases could have attenuated associations, as participants with poorer lifestyle behaviors and lower HRQoL may have been less likely to participate. In addition, the survey completion rate was relatively low (32%) and the sample was skewed towards women, which may limit the generalizability of these findings. Next, body composition was included as a lifestyle behavior since it aligns with one of the WCRF/AICR lifestyle recommendations. However, it may also be seen as a mediating factor, potentially influencing the validity of its findings. Finally, we cannot rule out the possibility of false positives due to multiple testing. Nevertheless, we focused on identifying trends and emphasizing the strength of associations rather focusing on statistical significance of individual associations.

### Future research directions

Many of the unhealthy lifestyle behaviors that AYA cancer survivors have could be addressed and ameliorated through interventions. Programs focusing on improving dietary habits, PA, and having a healthy body composition have shown notable success in promoting healthier lifestyle practices(49–51). However, these improvements have not consistently translated to better HRQoL and reduced fatigue. Given the preference for remote interventions among AYAs(50, 52), leveraging technology such as apps, wearable devices, and social networking platforms presents promising opportunities(53). These tools not only facilitate tracking and monitoring of behaviors, but also provide engagement and social support(54), which are critical for sustained lifestyle changes. Moreover, integrating lifestyle-focused survivorship care into standard practice is essential, as it remains underdeveloped despite growing evidence on the long-term and late effects experienced by AYA cancer survivors. Beyond treatment-focused care, comprehensive survivorship programs are necessary to address ongoing health challenges. Additionally, exploring the determinants of lifestyle behaviors over time will help pinpoint critical periods for intervention. Finally, evaluating the impact of interventions initiated during cancer treatment or immediately following diagnosis may uncover strategies for to instill long-term behavioral changes. Such approaches could play a preventive role in mitigating late effects and ensure sustained improvements in HRQoL.

### Conclusion

This study underscores the need to improve adherence to lifestyle recommendations among long-term AYA cancer survivors. Addressing gaps in having a healthy body composition, PA and nutrition can empower survivors to achieve better health outcomes. The findings from this study provide a foundation for developing interventions that not only enhance physical functioning, but also improve HRQoL and fatigue outcomes in this unique AYA cancer survivor population.

## Data Availability

All data produced in the present study are available upon reasonable request to the authors

## Statements and declarations

### Author contributions

Carla Vlooswijk, Olga Husson and Sandra Beijer contributed to the study conception and design. Material preparation and data collection was performed by Carla Vlooswijk and Olga Husson. Data analyses were done by Marlou Floor Kenkhuis. The first draft of the manuscript was written by Marlou Floor Kenkhuis and Laurien Buffart and all authors commented on previous versions of the manuscript. All authors read and approved the final manuscript.

### Ethics approval

The SURVAYA study was conducted according to the Declaration of Helsinki guidelines, and approved by the NKI Institutional Review Board (IRB-IRBd18122).

### Consent to participate

Informed consent was obtained from all subjects (responders) involved in the study.

## Acknowledgments

The authors wish to thank all the AYA cancer survivors for their participation in the study, the registration teams of the Netherlands Comprehensive Cancer Organisation (IKNL) for the collection of data for the Netherlands Cancer Registry and the PROFILES registry for distribution and handling of the questionnaires.

## References

1. Adolescent and Young Adult Oncology Progress review Group. Closing the gap: research and care imperatives for adolescents and young adults with cancer. 2006.

2. Lewis DR, Seibel NL, Smith AW, Stedman MR. Adolescent and young adult cancer survival. J Natl Cancer Inst Monogr. 2014;2014(49):228–35.

3. Ferrari A, Stark D, Peccatori FA, Fern L, Laurence V, Gaspar N, et al. Adolescents and young adults (AYA) with cancer: a position paper from the AYA Working Group of the European Society for Medical Oncology (ESMO) and the European Society for Paediatric Oncology (SIOPE). ESMO Open. 2021;6(2):100096.

4. Wen YF, Chen MX, Yin G, Lin R, Zhong YJ, Dong QQ, Wong HM. The global, regional, and national burden of cancer among adolescents and young adults in 204 countries and territories, 1990–2019: a population-based study. Journal of Hematology & Oncology. 2021;14(1):89.

5. Bleyer A, Ferrari A, Whelan J, Barr RD. Global assessment of cancer incidence and survival in adolescents and young adults. Pediatr Blood Cancer. 2017;64(9).

6. van der Meer DJ, Karim-Kos HE, van der Mark M, Aben KKH, Bijlsma RM, Rijneveld AW, et al. Incidence, Survival, and Mortality Trends of Cancers Diagnosed in Adolescents and Young Adults (15-39 Years): A Population-Based Study in The Netherlands 1990-2016. Cancers (Basel). 2020;12(11).

7. Morton LM, Onel K, Curtis RE, Hungate EA, Armstrong GT. The rising incidence of second cancers: patterns of occurrence and identification of risk factors for children and adults. Am Soc Clin Oncol Educ Book. 2014:e57–67.

8. Langeveld N, Ubbink M, Smets E, Group DLES. ‘I don’t have any energy’: the experience of fatigue in young adult survivors of childhood cancer. European Journal of Oncology Nursing. 2000;4(1):20–8.

9. Forbes C, Tanner S, Engstrom T, Lee WR, Patel D, Walker R, et al. Patient Reported Fatigue Among Adolescent and Young Adult Cancer Patients Compared to Non-Cancer Patients: A Systematic Review and Meta-Analysis. Journal of Adolescent and Young Adult Oncology. 2024;13(2):242–50.

10. Poort H, Kaal SE, Knoop H, Jansen R, Prins JB, Manten-Horst E, et al. Prevalence and impact of severe fatigue in adolescent and young adult cancer patients in comparison with population-based controls. Supportive Care in Cancer. 2017;25:2911–8.

11. van Zutphen M, Kampman E, Giovannucci EL, van Duijnhoven FJ. Lifestyle after colorectal cancer diagnosis in relation to survival and recurrence: a review of the literature. Current colorectal cancer reports. 2017;13:370–401.

12. Gopalakrishna A, Longo TA, Fantony JJ, Van Noord M, Inman BA. Lifestyle factors and health-related quality of life in bladder cancer survivors: a systematic review. Journal of Cancer Survivorship. 2016;10:874–82.

13. Blanchard CM, Courneya KS, Stein K. Cancer survivors’ adherence to lifestyle behavior recommendations and associations with health-related quality of life: results from the American Cancer Society’s SCS-II. Journal of Clinical Oncology. 2008;26(13):2198–204.

14. Wurz A, Brunet J. The effects of physical activity on health and quality of life in adolescent cancer survivors: a systematic review. JMIR cancer. 2016;2(1):e5431.

15. Munsie C, Ebert J, Joske D, Ackland T. The benefit of physical activity in adolescent and young adult cancer patients during and after treatment: a systematic review. Journal of adolescent and young adult oncology. 2019;8(5):512–24.

16. Zhi X, Xie M, Zeng Y, Liu J-e, Cheng AS. Effects of exercise intervention on quality of life in adolescent and young adult cancer patients and survivors: a meta-analysis. Integrative cancer therapies. 2019;18:1534735419895590.

17. Demark-Wahnefried W, Case LD, Blackwell K, Marcom PK, Kraus W, Aziz N, et al. Results of a diet/exercise feasibility trial to prevent adverse body composition change in breast cancer patients on adjuvant chemotherapy. Clinical breast cancer. 2008;8(1):70–9.

18. Quidde J, von Grundherr J, Koch B, Bokemeyer C, Escherich G, Valentini L, et al. Improved nutrition in adolescents and young adults after childhood cancer-INAYA study. BMC cancer. 2016;16:1–7.

19. Carla Vlooswijk LVvdP-F, Silvie H. M. Janssen, Esther Derksen, Milou J. P. Reuvers, Rhodé Bijlsma, Suzanne E. J. Kaal, Jan Martijn Kerst, Jacqueline M. Tromp, Monique E. M. M. Bos, Tom van der Hulle, Roy I. Lalisang, Janine Nuver, Mathilde C. M. Kouwenhoven, Winette T. A. van der Graaf and Olga Husson. Recruiting Adolescent and Young Adult Cancer Survivors for Patient-Reported Outcome Research: Experiences and Sample Characteristics of the SURVAYA Study. Current Oncology. 2022;29(8).

20. van de Poll-Franse LV, Horevoorts N, van Eenbergen M, Denollet J, Roukema JA, Aaronson NK, et al. The Patient Reported Outcomes Following Initial treatment and Long term Evaluation of Survivorship registry: scope, rationale and design of an infrastructure for the study of physical and psychosocial outcomes in cancer survivorship cohorts. Eur J Cancer. 2011;47(14):2188–94.

21. Shams-White MM, Brockton NT, Mitrou P, Romaguera D, Brown S, Bender A, et al. Operationalizing the 2018 World Cancer Research Fund/American Institute for Cancer Research (WCRF/AICR) Cancer Prevention Recommendations: A Standardized Scoring System. Nutrients. 2019;11(7).

22. Kenkhuis M-F, Mols F, van Roekel EH, Breedveld-Peters JJ, Breukink SO, Janssen-Heijnen ML, et al. Longitudinal associations of adherence to the World Cancer Research Fund/American Institute for Cancer Research (WCRF/AICR) lifestyle recommendations with quality of life and symptoms in colorectal cancer survivors up to 24 months post-treatment. Cancers. 2022;14(2):417.

23. Vidra N, Beeren I, van Zutphen M, Aben KK, Kampman E, Witjes JA, et al. Longitudinal associations of adherence to lifestyle recommendations and health-related quality of life in patients with non-muscle invasive bladder cancer. International Journal of Cancer. 2023;152(10):2032–42.

24. Allison P. Alcohol consumption is associated with improved health-related quality of life in head and neck cancer patients. Oral oncology. 2002;38(1):81–6.

25. World Cancer Research Fund/American Institute for Cancer Research, editor. Continuous Update Project Expert Report 2018. Recommendations and public health and policy implications. Available at dietandcancerreport.org. 2018.

26. Pols MA, Peeters PH, Ocké MC, Slimani N, Bueno-de-Mesquita HB, Collette HJ. Estimation of reproducibility and relative validity of the questions included in the EPIC Physical Activity Questionnaire. Int J Epidemiol. 1997;26 Suppl 1:S181–9.

27. Ainsworth BE, Haskell WL, Leon AS, Jacobs DR, Jr., Montoye HJ, Sallis JF, Paffenbarger RS, Jr. Compendium of physical activities: classification of energy costs of human physical activities. Med Sci Sports Exerc. 1993;25(1):71–80.

28. Ainsworth BE, Haskell WL, Whitt MC, Irwin ML, Swartz AM, Strath SJ, et al. Compendium of physical activities: an update of activity codes and MET intensities. Med Sci Sports Exerc. 2000;32(9 Suppl):S498–504.

29. Vlooswijk C, Oerlemans S, Ezendam NPM, Schep G, Slot S, Thong MSY, et al. Physical Activity is Associated with Health Related Quality of Life in Lymphoma Survivors Regardless of Body Mass Index; Results from the Profiles Registry. Nutr Cancer. 2022;74(1):158–67.

30. Gavioli C, Vlooswijk C, Janssen SH, Kaal SE, Kerst JM, Tromp JM, et al. Adherence to the World Cancer Research Fund/American Institute for Cancer Research recommendations for cancer prevention in adolescent and young adult (AYA) cancer survivors: results from the SURVAYA study. Journal of Cancer Survivorship. 2024:1–15.

31. Center for Disease Control and Prevention (CDC). Healthy Weight: Assessing Your Weight [Available from: https://www.cdc.gov/healthyweight/assessing/index.html.

32. National Heart Lung and Blood Institute. Assessing Your Weight and Health Risk [Available from: https://www.nhlbi.nih.gov/health/educational/lose_wt/risk.htm.

33. Fritz A, Percy, Constance, Jack, Andrew, Shanmugaratnam, Kanagaratnam, Sobin, Leslie H. et al. International classification of diseases for oncology. 3rd ed. ed2000.

34. Sobin L, Gospodarowicz MKW, C. TNM classification of malignant tumours. New York: Wiley; 2011.

35. Breedveld-Peters JJ, Koole JL, Müller-Schulte E, van der Linden BW, Windhausen C, Bours MJ, et al. Colorectal cancers survivors’ adherence to lifestyle recommendations and cross-sectional associations with health-related quality of life. British Journal of Nutrition. 2018;120(2):188–97.

36. van Zutphen M, Hof JP, Aben KK, Kampman E, Witjes JA, Kiemeney LA, Vrieling A. Adherence to lifestyle recommendations after non-muscle invasive bladder cancer diagnosis and risk of recurrence. The American Journal of Clinical Nutrition. 2023;117(4):681–90.

37. Winkels RM, van Lee L, Beijer S, Bours MJ, van Duijnhoven FJ, Geelen A, et al. Adherence to the World Cancer Research Fund/American Institute for Cancer Research lifestyle recommendations in colorectal cancer survivors: results of the PROFILES registry. Cancer medicine. 2016;5(9):2587–95.

38. Malcomson FC, Parra-Soto S, Ho FK, Lu L, Celis-Morales C, Sharp L, Mathers JC. Adherence to the 2018 World Cancer Research Fund (WCRF)/American Institute for Cancer Research (AICR) Cancer Prevention Recommendations and risk of 14 lifestyle-related cancers in the UK Biobank prospective cohort study. BMC medicine. 2023;21(1):407.

39. Nolte S, Liegl G, Petersen M, Aaronson N, Costantini A, Fayers P, et al. General population normative data for the EORTC QLQ-C30 health-related quality of life questionnaire based on 15,386 persons across 13 European countries, Canada and the Unites States. European journal of cancer. 2019;107:153–63.

40. Zhang Y, Kesler SR, Dietrich J, Chao HH. Cancer-Related Cognitive Impairment: A Practical Guide for Oncologists. JCO Oncology Practice. 2025:OP-24-00953.

41. Von Ah D. Evidence-based interventions for cancer-and treatment-related cognitive impairment. Number 6/December 2014. 2014;18(6):17–25.

42. Haines ER, Lux L, Smitherman AB, Kessler ML, Schonberg J, Dopp A, et al. An actionable needs assessment for adolescents and young adults with cancer: the AYA Needs Assessment & Service Bridge (NA-SB). Supportive Care in Cancer. 2021;29:4693–704.

43. Schleicher E, McAuley E, Courneya KS, Anton P, Ehlers DK, Phillips SM, et al. Moderators of physical activity and quality of life response to a physical activity intervention for breast cancer survivors. Supportive Care in Cancer. 2023;31(1):53.

44. Russo S, Cinausero M, Gerratana L, Bozza C, Iacono D, Driol P, et al. Factors affecting patient’s perception of anticancer treatments side-effects: an observational study. Expert opinion on drug safety. 2014;13(2):139–50.

45. Jansen S, Otten W, Van de Velde C, Nortier J, Stiggelbout A. The impact of the perception of treatment choice on satisfaction with treatment, experienced chemotherapy burden and current quality of life. British Journal of Cancer. 2004;91(1):56–61.

46. Silva SM, Moreira HC, Canavarro MC. Examining the links between perceived impact of breast cancer and psychosocial adjustment: the buffering role of posttraumatic growth. Psycho-Oncology. 2012;21(4):409–18.

47. Olfert MD, Barr ML, Charlier CM, Famodu OA, Zhou W, Mathews AE, et al. Self-reported vs. measured height, weight, and BMI in young adults. International journal of environmental research and public health. 2018;15(10):2216.

48. ECF - European Cyclists’ Federation. [

49. Pugh G, Gravestock HL, Hough RE, King WM, Wardle J, Fisher A. Health Behavior Change Interventions for Teenage and Young Adult Cancer Survivors: A Systematic Review. J Adolesc Young Adult Oncol. 2016;5(2):91–105.

50. Skiba MB, McElfresh JJ, Howe CL, Crane TE, Kopp LM, Jacobs ET, Thomson CA. Dietary Interventions for Adult Survivors of Adolescent and Young Adult Cancers: A Systematic Review and Narrative Synthesis. J Adolesc Young Adult Oncol. 2020;9(3):315–27.

51. Munsie C, Ebert J, Joske D, Ackland T. A randomised controlled trial investigating the ability for supervised exercise to reduce treatment-related decline in adolescent and young adult cancer patients. Support Care Cancer. 2022;30(10):8159–71.

52. Carretier J, Boyle H, Duval S, Philip T, Laurence V, Stark DP, et al. A Review of Health Behaviors in Childhood and Adolescent Cancer Survivors: Toward Prevention of Second Primary Cancer. J Adolesc Young Adult Oncol. 2016;5(2):78–90.

53. Lubans DR, Morgan PJ, Tudor-Locke C. A systematic review of studies using pedometers to promote physical activity among youth. Prev Med. 2009;48(4):307–15.

54. West JH, Hall PC, Hanson CL, Barnes MD, Giraud-Carrier C, Barrett J. There’s an app for that: content analysis of paid health and fitness apps. J Med Internet Res. 2012;14(3):e72.

